# Germline Variants Absent from Global Reference Databases in Malawian DLBCL Patients: Implications for Variant Interpretation in African Genomic Medicine

**DOI:** 10.64898/2026.09.10.26362726

**Authors:** Kelvin Chidothi, Samuel Gwayi, Benjamin Kumwenda

## Abstract

**Background:** African populations remain underrepresented in global genomic reference databases, complicating germline variant interpretation in clinical and research settings. This concern is particularly relevant in Malawi, where population-matched allele-frequency resources are limited. We assessed germline variants in Malawian patients with diffuse large B-cell lymphoma (DLBCL) that were absent from commonly used global reference databases.

**Methods:** Whole-exome sequencing data from 55 histologically confirmed DLBCL cases recruited at Kamuzu Central Hospital, Lilongwe, were analyzed. Germline variants were called using GATK HaplotypeCaller and annotated with Ensembl VEP v110. Cohort allele frequencies were compared with gnomAD v3.1.2, ExAC v1.0, and the 1000 Genomes Project Phase 3 using African (AFR) and European (EUR) reference subsets. Variants absent from all three databases were designated database-absent variants (DAVs); this designation indicates absence from the queried reference panels and does not establish population restriction. Functional consequence, genomic distribution, and enrichment in immune-related loci were evaluated.

**Results:** We identified 127,456 PASS-filtered germline variants across 55 samples. Of these, 332 variants (0.26%) were absent from gnomAD, ExAC, and 1000 Genomes and were distributed across 289 genes (median, one variant per gene; range, 1-8). Predicted loss-of-function variants comprised 16.9% of DAVs compared with 7.8% of the full call set (odds ratio, 2.41; 95% confidence interval, 1.79-3.24; *p* < 0.001). Seventy-eight DAVs (23.5%) occurred in HLA class I loci (HLA-A, HLA-B, and HLA-C; Fisher’s exact test, *p* < 0.001). Allele-frequency distributions differed from both AFR and EUR reference populations, with greater divergence from EUR (Kolmogorov-Smirnov *D* = 0.31) than AFR (*D* = 0.13). The cohort also showed an increased proportion of low-frequency variants relative to both references.

**Conclusions:** Database-absent germline variants were observed in this Malawian DLBCL cohort, including variants with predicted loss-of-function consequences and variants in HLA class I loci. These findings highlight gaps in the representation of Malawian and Southern African genomes in commonly used reference resources and the resulting potential for uncertainty in variant interpretation. Because this case-only cohort lacked matched population controls and sequencing depth was limited, DAVs should not be interpreted as definitively Malawian-specific or pathogenic. Larger, well-covered studies including matched Malawian and Southern African controls are needed to establish population frequencies and improve equitable genomic interpretation.

## 1. Introduction

The accurate clinical interpretation of germline genetic variants depends fundamentally on the availability of population-matched allele frequency data. A variant classified as pathogenic in one population may be a common benign polymorphism in another, and a variant absent from reference databases may represent either a genuinely novel disease-causing mutation or simply a gap in population sampling. This distinction carries direct clinical consequences: erroneous pathogenicity classifications contribute to misdiagnosis, inappropriate therapeutic decisions, and inequitable access to precision medicine [1,2].

Despite this, global genomic reference databases remain heavily skewed toward populations of European and East Asian ancestry. The Genome Aggregation Database (gnomAD), the most widely used frequency reference, contains over 125,000 exomes and 76,000 genomes, yet African ancestry individuals represent less than 12% of the total sample, and representation from sub-Saharan African countries, including Malawi, Zambia, Mozambique, and Zimbabwe, is negligible or absent [3]. The 1000 Genomes Project Phase 3 includes 661 individuals from African populations, but these are drawn predominantly from West African (Yoruba, Ewe, Mende) and East African (Luhya, Maasai) groups [4]. Southern African populations, which carry distinct genetic signatures reflecting ancient population structure and admixture [20], are systematically absent from these catalogues [5].

Africa harbors the greatest human genetic diversity on earth, a consequence of the continent’s role as the geographic origin of anatomically modern humans and its complex population history of migration, isolation, and admixture [6]. African populations therefore carry the largest burden of population-specific rare variants, alleles present at appreciable frequency locally but absent or vanishingly rare in non-African populations. When these variants are encountered in clinical genomic testing and compared against databases lacking African representation, they are disproportionately flagged as variants of uncertain significance (VUS) or worse, misclassified as pathogenic [7,8]. This phenomenon has been documented across cancer predisposition genes, cardiovascular pharmacogenomics, and rare Mendelian disease, and represents a systematic disparity in the quality of genomic medicine available to African patients relative to patients of European ancestry [9,10].

In Malawi, the problem is compounded by an acute cancer burden. Lymphomas account for approximately 46% of all cancer diagnoses, and Diffuse Large B-Cell Lymphoma (DLBCL), the most common subtype, presents at a median age nearly five decades younger than in Western populations, with five-year survival below 30% [11,12]. HIV co-morbidity, endemic Epstein-Barr virus exposure, and limited access to molecular diagnostics create a disease context fundamentally different from that informing existing genomic tools and classification frameworks [13]. Yet no population-specific germline variant catalogue has been generated for any Malawian cancer cohort, leaving clinicians and researchers entirely reliant on reference data derived from populations with profoundly different genetic backgrounds.

This study addresses this gap directly. Using whole-exome sequencing of 55 histologically confirmed DLBCL cases from Kamuzu Central Hospital, we characterize germline variants absent from all three major global reference databases, gnomAD, ExAC, and the 1000 Genomes Project. We describe their functional class distribution, genomic localization, and enrichment patterns, with particular attention to immune-related loci and loss-of-function consequences that carry the greatest clinical interpretive risk. We situate these findings within the broader context of genomic equity in Africa and articulate the minimum data infrastructure required to enable accurate variant interpretation in Malawian patients.

## 2. Materials and Methods

### 2.1 Study Design and Cohort

This study employed a cross-sectional design using whole-exome sequencing data from 55 histologically confirmed DLBCL cases recruited from the Kamuzu Central Hospital (KCH) Lymphoma Study, a prospective observational cohort initiated in June 2013. Diagnoses were confirmed through immunohistochemistry and interdisciplinary telepathology review. Of 77 DLBCL patients in the parent cohort, 55 had whole-exome sequencing data available. All 55 were included; no additional exclusion criteria were applied beyond those of the original study.

Ethical approval was obtained from the University of North Carolina Institutional Review Board (UNC IRB 12-2255), the National Health Science Research Committee of Malawi (NHSRC 1107), and the College of Medicine Research and Ethics Committee (COMREC, Reference ID: P.07/25-1690). All data were de-identified prior to analysis.

Demographic and clinical characteristics were available at the level of the parent KCH Lymphoma Study cohort (n = 77), from which the 55 sequenced cases were drawn: 45 males and 32 females, with a median age of 46 years (range, 22-80 years). Fifty-four patients (70.1%) were HIV-positive, of whom 35 had received antiretroviral therapy for at least 6 months and 18 had not (ART status missing for one patient); 23 patients (29.9%) were HIV-negative. Patient-level demographic and clinical metadata could not be linked to the specific 55-patient whole-exome sequencing subset analyzed here, because this secondary analysis used de-identified genomic data only; the characteristics above therefore describe the parent cohort from which the sequenced subset was drawn rather than the analytic subset itself.

### 2.2 Sequencing and Variant Calling

Raw FASTQ files were quality-assessed using FastQC v0.12.1 and MultiQC. Adapter trimming and quality filtering used Trimmomatic v0.39 (TruSeq3 adapters; Phred threshold Q30; minimum read length 36 bases). Trimmed reads were aligned to GRCh38 using BWA-MEM v0.7.17. BAM files were processed per GATK v4 best practices, including duplicate marking, read group assignment, and base quality score recalibration (BQSR) using dbSNP and Mills known variant sites.

Germline variant discovery used GATK HaplotypeCaller in GVCF mode; per-sample GVCFs were consolidated with GenomicsDBImport and jointly genotyped using GenotypeGVCFs. SNPs and INDELs were filtered using CNN-based variant scoring (CNNScoreVariants) at tranche thresholds of 99.95% and 99.4% respectively; only PASS-filtered variants were retained.

Functional annotation used Ensembl VEP v110 (GRCh38 offline mode), incorporating SIFT, PolyPhen-2, CADD, gnomAD v3.1.2 allele frequencies, and ClinVar annotations.

The analysis used existing whole-exome sequencing data from the Kamuzu Central Hospital Lymphoma Study. The available dataset was generated at low sequencing coverage, with a mean read depth of 3.5x, and was therefore used for exploratory, cohort-level analysis rather than clinical-grade interpretation of individual variants. To reduce technical artefacts, reads underwent quality assessment, adapter trimming, alignment to GRCh38, duplicate marking, and base-quality score recalibration. Variants were jointly genotyped and retained only when they passed CNN-based tranche filtering; calls in low-complexity regions and segmental duplications were excluded from the database-absent variant analysis. In this manuscript, “PASS-filtered” denotes retention after these computational filters and should not be interpreted as orthogonal validation of individual rare variants.

### 2.3 African Ancestry Verification

African ancestry was verified by principal component analysis (PCA) merging study samples with the 1000 Genomes Project Phase 3 reference populations, following LD pruning (window 50 SNPs, step 10, r^2^ threshold 0.1) and MAF filtering (MAF > 0.05), performed using PLINK v1.9.

### 2.4 Population-Specific Variant Identification

Reference populations were selected using the ancestry labels provided by each database. For 1000 Genomes Phase 3, the AFR comparison consisted of seven subpopulations: four from West Africa (YRI, ESN, GWD, MSL), one from East Africa (LWK), and two African-diaspora populations (ASW, ACB); no Malawian or other Southern African reference population was included. For gnomAD and ExAC, comparisons used their available broad African/African American and European-derived ancestry strata. Sample counts represent release-level totals; the allele number available for a specific variant may be lower because of locus-level coverage and quality-control filtering.

Variants were compared with gnomAD v3.1.2, ExAC v1.0, and the 1000 Genomes Project Phase 3 using ancestry-stratified African (AFR) and European (EUR) reference subsets. The three reference resources differ in sequencing technology, sample composition, genome build, and quality-control procedures; database absence was therefore interpreted as absence from the queried resources rather than definitive evidence of novelty or population restriction. gnomAD v3.1.2 is a whole-genome release, whereas its most recent exome release was gnomAD v2.1. ExAC was retained as a supplementary exome-based comparison resource because it may contain exome observations not represented in the gnomAD v3.1.2 genome dataset.

ExAC v1.0 was included as a legacy supplementary reference alongside gnomAD v3.1.2 and the 1000 Genomes Project Phase 3. Given substantial overlap between ExAC and gnomAD, gnomAD was treated as the primary contemporary population-frequency resource, while ExAC served only as a conservative historical cross-check. A variant was classified as database-absent when no exact allele-level match was identified in any of the three resources.

Variants not observed in any of the three databases at any reported allele frequency were designated database-absent variants (DAVs), after excluding calls in low-complexity regions and segmental duplications. This designation indicates absence from the queried reference panels and does not establish that a variant is restricted to the Malawian population or to any other population. Definitive assessment of population specificity would require comparison with appropriately matched Malawian or Southern African population controls, which were not available in this study.

### 2.5 Functional Classification and Enrichment Analysis

Enrichment of LoF variants among database-absent variants relative to the full call set was assessed by Fisher’s exact test with odds-ratio estimation. Genomic enrichment of database-absent variants in HLA class I loci (HLA-A, HLA-B, and HLA-C) was assessed against genome-wide expectation by Fisher’s exact test. ClinVar annotation was used to describe previously reported variant annotations; it was not used to establish population specificity.

## 3. Results

### 3.1 Sequencing Quality

All 55 samples met the predefined quality-control thresholds and were retained for downstream analysis. Mean sequencing depth was 3.5 +/- 0.8x (range, 2.1-5.3x), with a mean mapping rate of 96.3% +/- 2.1%, 94.7% +/- 1.8% properly paired reads, a mean duplication rate of 12.1% +/- 3.4% and mean base quality of Q36.8 +/- 1.2. Target-region coverage at >=10x was 78.3% +/- 6.2%. Following quality control and filtering, 127,456 germline variants were retained across the cohort, comprising 113,824 single-nucleotide variants (89.3%), 7,234 insertions (5.7%), and 6,398 deletions (5.0%). The mean number of retained variants per sample was 2,317 (range, 1,856-2,891). The transition/transversion ratio was 2.08 +/- 0.03, consistent with the expected range for exome sequencing data and providing an additional quality check for the overall call set. Nevertheless, the low mean sequencing depth warrants cautious interpretation of individual rare variants, particularly those at lower-coverage loci.

#### 3.1.1 Coverage and Genotype Quality at Database-Absent Variant Loci

Across the cohort, 78.3% +/- 6.2% of target regions achieved coverage of >=10x, despite a mean sequencing depth of 3.5x +/- 0.8x. However, site-specific coverage, genotype quality, and allele-balance metrics were not available for the 332 database-absent variants. The confidence of individual DAV calls, particularly those at lower-coverage loci, should therefore be interpreted cautiously and requires confirmation through targeted or higher-coverage sequencing.

### 3.2 Allele Frequency Distribution and Ancestry Validation

PCA confirmed that all 55 samples clustered within the African superpopulation reference, validating the appropriateness of African ancestry calibration. Allele frequency distributions in the Malawian cohort differed significantly from both the African (AFR) and European (EUR) superpopulations of the 1000 Genomes Project Phase 3 (chi-square test across all MAF bins: χ^2^ = 4,523.7, p < 0.001).

**Figure 1:**
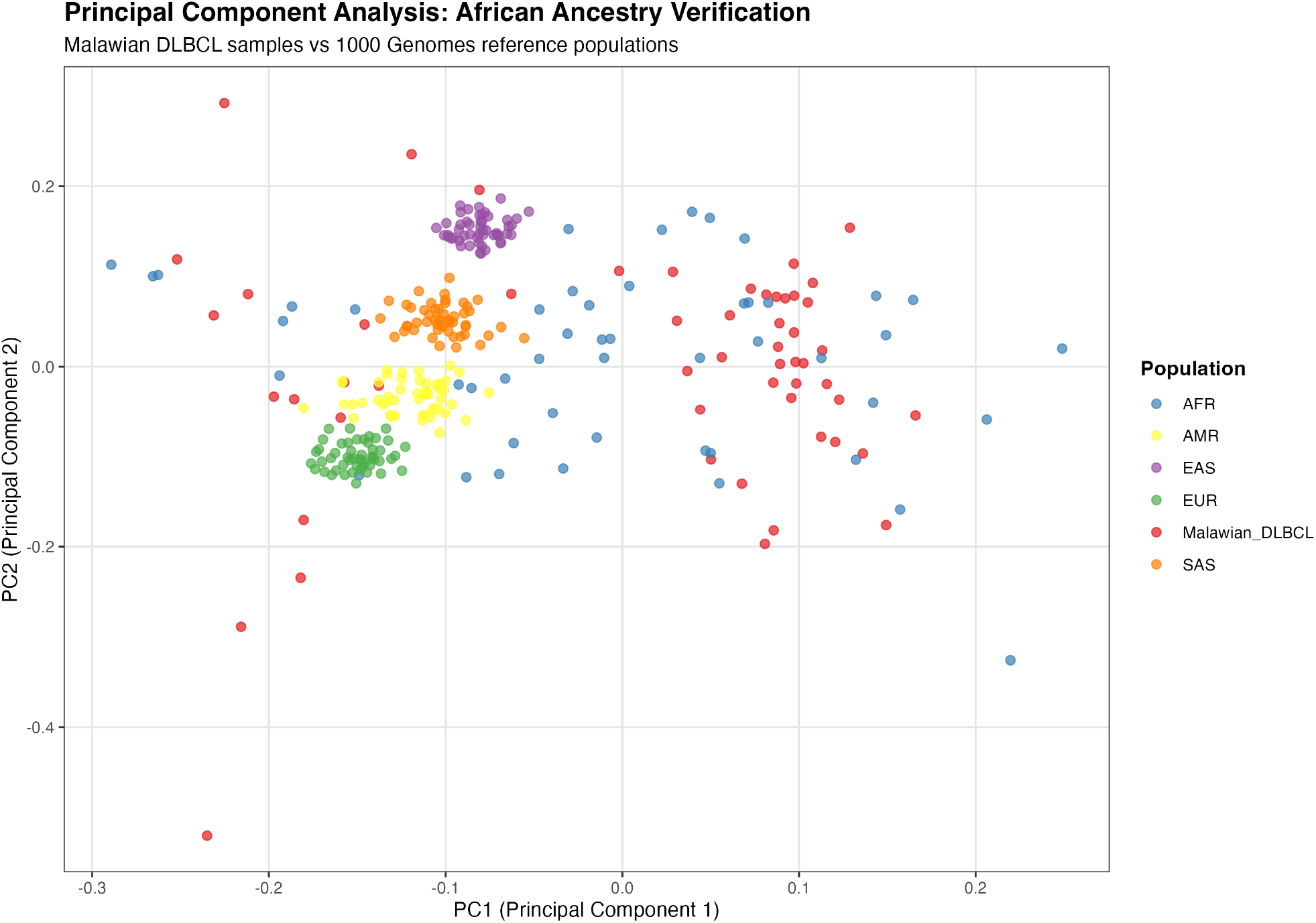
Principal component analysis of Malawian DLBCL samples and 1000 Genomes reference populations. The Malawian DLBCL samples cluster predominantly with the African-ancestry reference population (AFR), supporting African ancestry assignment for downstream analyses. Each point represents one individual; colors indicate reference population or study cohort.

Kolmogorov–Smirnov analysis confirmed closer alignment with the African reference (D = 0.13, p = 0.003) than with the European reference (D = 0.31, p < 0.001), yet the two distributions were still significantly different, reflecting population-specific diversity not captured by any existing reference panel.

The 1000 Genomes AFR comparator comprised predominantly West African populations (YRI, ESN, GWD, and MSL), one East African population (LWK), and African-diaspora populations (ASW and ACB), with no Malawian or Southern African population represented (Table 1).

**Table 1.**
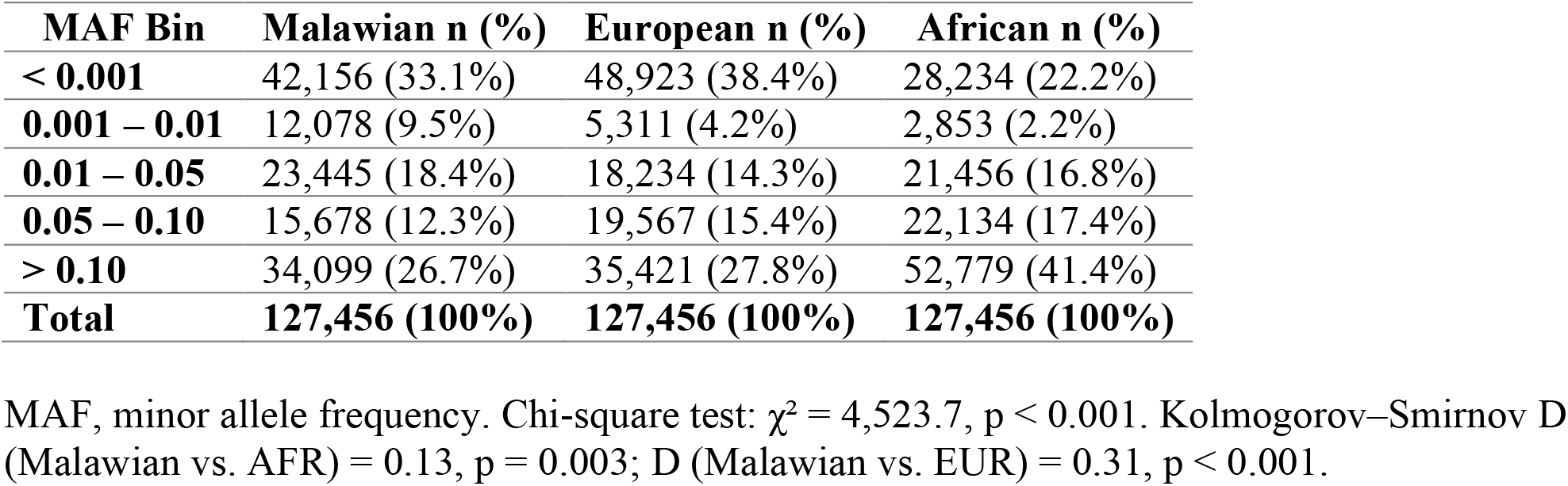
Allele frequency distribution across the Malawian cohort and global reference populations.

| MAF Bin | Malawian n (%) | European n (%) | African n (%) |
| --- | --- | --- | --- |
| < 0.001 | 42,156 (33.1%) | 48,923 (38.4%) | 28,234 (22.2%) |
| 0.001 – 0.01 | 12,078 (9.5%) | 5,311 (4.2%) | 2,853 (2.2%) |
| 0.01 – 0.05 | 23,445 (18.4%) | 18,234 (14.3%) | 21,456 (16.8%) |
| 0.05 – 0.10 | 15,678 (12.3%) | 19,567 (15.4%) | 22,134 (17.4%) |
| > 0.10 | 34,099 (26.7%) | 35,421 (27.8%) | 52,779 (41.4%) |
| **Total** | **127,456 (100%)** | **127,456 (100%)** | **127,456 (100%)** |

MAF, minor allele frequency. Chi-square test: $\chi^2 = 4,523.7$ , $p < 0.001$ . Kolmogorov–Smirnov D (Malawian vs. AFR) = 0.13, $p = 0.003$ ; D (Malawian vs. EUR) = 0.31, $p < 0.001$ .

Subpopulation sample sizes were 108 (YRI), 99 (ESN), 113 (GWD), 85 (MSL), 99 (LWK), 61 (ASW), and 96 (ACB), totaling 661 African-ancestry individuals [4].

The Malawian cohort showed notable enrichment of low-frequency variants (MAF 0.001–0.01) at 9.5%, compared to 4.2% in Europeans and 2.2% in the African reference panel. This enrichment is consistent with population-specific rare variation that is underrepresented even in African superpopulation references, which are dominated by West African samples. Variants with MAF > 0.10 were present at 26.7% in the Malawian cohort versus 41.4% in the African reference, reflecting the expected depletion of common variants when Malawian-specific alleles are compared against a broader African reference (Table 1).

### 3.3 Population-Specific Variants Absent from Global Databases

In the filtered variant set, 332 variants (0.26%) had no allele-level match in gnomAD v3.1.2, ExAC v1.0, or the 1000 Genomes Project Phase 3 and were classified as database-absent variants (DAVs). ExAC was retained as a conservative legacy cross-reference; however, because it substantially overlaps with and has been superseded by gnomAD, it was not interpreted as an independent population reference. Database absence should therefore not be interpreted as evidence that a variant is population-specific, Malawi-specific, or pathogenic.

The 332 DAVs were distributed across 289 genes, with a median of one variant per gene (range, 1-8), indicating broad genomic dispersion rather than concentration at a small number of loci. Of these variants, 201 (60.5%) occurred in genes with ClinVar records containing African-ancestry-related annotations, whereas 131 (39.5%) occurred in genes without such annotations. These findings describe the coverage and annotation limitations of available reference resources rather than evidence of disease association or population specificity.

### 3.4 Functional Class Distribution of Database-Absent Variants

Loss-of-function variants were significantly enriched among database-absent variants relative to the genome-wide call set. Combined LoF classes (frameshift, nonsense, splice-site) comprised 16.9% of database-absent variants, compared to 7.8% genome-wide (OR = 2.41, 95% CI: 1.79– 3.24, p < 0.001). Among individual LoF classes, nonsense variants showed the strongest enrichment (enrichment ratio 3.18), followed by frameshift variants (1.92) and splice-site variants (1.80). Missense variants showed modest enrichment (ratio 1.07), while synonymous variants were slightly under-represented (ratio 0.89) (Table 2).

**Table 2.** Functional class distribution of database-absent variants versus the full call set.

| **Functional Class** | **Database-absent variants n (%)** | **All Variants n (%)** | **Enrichment Ratio** |
| --- | --- | --- | --- |
| **Missense** | 187 (56.3%) | 67,234 (52.7%) | 1.07 |
| **Synonymous** | 89 (26.8%) | 38,456 (30.2%) | 0.89 |
| **Frameshift** | 23 (6.9%) | 4,567 (3.6%) | 1.92 |
| **Nonsense** | 18 (5.4%) | 2,134 (1.7%) | 3.18 |
| **Splice site** | 15 (4.5%) | 3,234 (2.5%) | 1.80 |
| **Total** | **332 (100%)** | **127,456 (100%)** | — |

Loss-of-function (LoF) variants comprise frameshift, nonsense, and splice-site classes: 16.9% of database-absent variants versus 7.8% genome-wide; OR = 2.41, 95% CI: 1.79–3.24, $p < 0.001$ . Enrichment ratio = proportion in database-absent variants ÷ proportion in all variants.

The synonymous under-representation among database-absent variants is mechanistically interpretable: synonymous variants are subject to weaker purifying selection than LoF variants and are therefore more likely to drift to higher frequencies at which they would be detected in existing African reference panels. LoF variants, by contrast, are subject to stronger purifying selection, maintaining them at low frequencies in all populations, and their enrichment among database-absent variants reflects the intersection of genuine novelty with high functional consequence.

### 3.5 Genomic Distribution: Concentration in HLA Class I Loci

A disproportionate concentration of database-absent variants was observed in immune-related loci. Seventy-eight variants (23.5% of all database-absent variants) were located within HLA class I genes; HLA-A, HLA-B, and HLA-C, representing highly significant enrichment relative to the expected genome-wide distribution (Fisher’s exact test, p < 0.001). Additional database-absent variants were concentrated in genes involved in antigen processing and presentation and cytokine signaling pathways, consistent with broader enrichment of immune-related loci.

The HLA region is the most polymorphic region in the human genome, and its diversity in African populations substantially exceeds that observed in European or East Asian cohorts [14]. This extraordinary polymorphism, driven by ancient balancing selection from infectious disease exposure, generates allele combinations in African populations that have no representation in current HLA reference databases. For DLBCL specifically, HLA class I loci have been consistently implicated as susceptibility regions in European and East Asian GWAS, yet their

African-specific allelic architecture has never been characterized in the context of lymphoma susceptibility [15].

## 4. Discussion

### 4.1 The Clinical Significance of Database-Absent Variants

The 332 database-absent variants identified in this cohort illustrate a practical challenge for germline variant interpretation when population-matched frequency data are unavailable.

Although these variants represented 0.26% of the PASS-filtered call set, they accounted for approximately six variants per exome. Their absence from gnomAD, ExAC, and 1000 Genomes indicates incomplete representation in these reference panels but does not establish that they are restricted to Malawian populations or associated with disease.

This limitation is relevant to use of the ACMG/AMP PM2 criterion, which considers absence or extreme rarity in population controls as supporting evidence in variant interpretation [16]. In populations that are poorly represented in reference databases, PM2 may be applied to benign alleles simply because population-frequency evidence is unavailable, a source of interpretive discordance documented even among experienced clinical laboratories applying ACMG/AMP criteria [17].

More critically, LoF variants were more than twice as prevalent among database-absent variants as in the genome-wide call set (OR = 2.41). This enrichment is precisely in the functional category that carries the greatest interpretive weight in clinical genomics: a nonsense or frameshift variant absent from all reference databases in a gene associated with cancer predisposition would receive strong pathogenic evidence under current guidelines, regardless of whether it is a neutral Malawian population-specific allele. The 39.5% of database-absent variants occurring in genes without any prior African allele documentation in ClinVar are particularly concerning, as these lack even the contextual signal that might temper erroneous pathogenic classification.

Loss-of-function annotation describes a predicted molecular consequence rather than clinical pathogenicity. Many predicted LoF variants are benign or have uncertain clinical significance; therefore, the observed enrichment should be interpreted as a potential source of variant-interpretation risk under current ACMG/AMP criteria, not as evidence that these specific variants are disease-causing.

While this enrichment may reflect genuine population-specific functional variation, alternative explanations should also be considered. Differential tolerance to loss-of-function variation across gene categories could contribute to this pattern independent of population-specific selection.

Annotation-based classification is itself a source of uncertainty: LoF status was assigned from VEP consequence annotation alone, without additional in-silico confidence filtering (e.g., LOFTEE) or experimental validation, so a subset of apparent LoF calls may reflect annotation artefacts rather than true protein-truncating variation. Technical factors related to the low sequencing depth used in this study may also differentially affect detection sensitivity across functional classes, and differences in the composition of the background (genome-wide) variant set between this cohort and the reference databases could further influence the observed enrichment ratio. These possibilities cannot be fully distinguished with the present data and should be considered alongside the population-genetic interpretation offered above.

### 4.2 HLA Diversity and Immune Gene Variation

The concentration of database-absent variants in HLA class I loci (23.5% of all database-absent variants; p < 0.001) reflects a well-established biological reality: HLA is the most polymorphic locus in the human genome precisely because pathogen-driven balancing selection has maintained extraordinary allelic diversity, and this diversity is greatest in African populations with the longest history of infectious disease co-evolution [18]. Sub-Saharan African HLA diversity includes numerous haplotypes entirely absent from European and Asian reference panels, and Malawi’s specific disease ecology, characterized by high HIV prevalence, endemic EBV and malaria, and schistosomiasis, has likely maintained distinctive HLA allele distributions through population-specific selection pressures [19].

For DLBCL, this matters directly. HLA class I associations have been among the most consistently replicated findings in GWAS conducted in European and East Asian populations, with specific alleles conferring relative risks of 1.2–1.5 for DLBCL susceptibility [15]. The mechanism, immune surveillance and antigen presentation of lymphoma-associated peptides, is biologically compelling and has implications for immunotherapy response as well as susceptibility. Yet the HLA alleles associated with DLBCL risk in European populations may not be the relevant variants in Malawian patients, who present with distinct disease epidemiology (earlier age of onset, higher HIV and EBV co-prevalence, different somatic mutation landscapes) and carry HLA haplotypes unrepresented in existing susceptibility studies. Our identification of 78 novel HLA class I variants absent from all major databases establishes that this diversity is real and currently uncatalogued.

An important technical consideration is that the HLA region has extreme polymorphism and substantial paralogous sequence structure, which can complicate short-read alignment and variant calling against a linear GRCh38 reference genome. The BWA-MEM-based workflow used in this study may therefore produce both false-positive and false-negative calls in this region, independently of genuine population variation. Accordingly, the 78 database-absent HLA class I variants identified here should be interpreted cautiously. HLA-specific approaches, including graph-based reference methods or dedicated callers such as HLA-LA or OptiType, were not used and would strengthen confidence in these findings in future validation studies.

### 4.3 Southern African Genetic Specificity

The observed differences between the Malawian cohort and available African reference populations should be interpreted considering the composition of those reference panels, which include predominantly West African, East African, and African-diaspora samples but no Malawian or other Southern African comparator population (Table 1).

The 131 database-absent variants occurring in genes without prior African-enriched ClinVar annotations may therefore reflect gaps in existing annotation and frequency resources. Their clinical interpretation will require larger population-based studies that establish allele frequencies in Malawian and related Southern African populations.

### 4.4 Implications for Variant Interpretation Frameworks

Frequency criteria supporting benign or pathogenic classification depend on adequately sampled reference populations. Population-matched genomic resources would improve the calibration of frequency-based interpretation in Malawian patients undergoing germline genomic testing and reduce reliance on absence from global databases as evidence of rarity.

Progress toward this goal is underway. Established initiatives, including H3Africa and the African Genome Variation Project, have expanded African genomic data generation across the continent [21,22], and more recent efforts, including the continued expansion of gnomAD with ancestry-aware frequency analyses and African-focused resources such as the African Genome Resource and emerging Southern African whole-genome datasets, demonstrate that broader sampling can improve reference panels. Clinically useful frequency catalogues specific to Malawi and neighbouring Southern African populations nonetheless remain limited, and the present case-only cohort is not itself a population reference panel; it identifies an evidence gap rather than filling it.

### 4.5 Limitations

Four limitations warrant explicit acknowledgment. First, the available whole-exome sequencing data had a mean depth of 3.5x, and 78.3% +/- 6.2% of target regions achieved coverage of >=10x. Although the analysis applied read-level quality control, joint genotyping, and PASS-only variant filtering, this depth is below that generally used for reliable clinical interpretation of individual rare variants. Low coverage can reduce sensitivity for heterozygous and rare variants and may increase uncertainty in individual calls. In addition, site-specific coverage, genotype quality, and allele-balance metrics were not available for the 332 database-absent variants; therefore, their confidence cannot be assessed uniformly at the individual-locus level. These variants should be regarded as exploratory findings requiring confirmation by targeted or higher-coverage sequencing, particularly for predicted loss-of-function and HLA-region variants.

Aggregate quality-control metrics for the PASS-filtered call set (mean mapping rate 96.3% +/- 2.1%; transition/transversion ratio 2.08 +/- 0.03) were within standard ranges for exome sequencing data, supporting the general reliability of the call set; however, these metrics were not stratified specifically for the 332 database-absent variants, and locus-level coverage and genotype-quality confirmation for these variants remains a priority for future validation.

Second, the cohort lacked healthy controls matched for Malawian ancestry. Consequently, the study could not determine whether database-absent variants are associated with DLBCL, are present in the broader Malawian population, or are shared with other Southern African populations. Matched population controls will be required to distinguish disease-associated variants from neutral population variation and to estimate population allele frequencies accurately.

Third, designation as a database-absent variant reflects absence from gnomAD, ExAC, and the 1000 Genomes Project at the time of analysis rather than confirmed novelty or population specificity. Reference databases continue to expand, and some of these variants may already be represented in newer releases or datasets not queried here.

Finally, the HLA region presents additional technical challenges because its extreme polymorphism and paralogous sequence structure can compromise short-read alignment and variant calling against a linear reference genome. HLA-specific calling approaches and orthogonal validation would be needed to strengthen confidence in the HLA findings.

### 5. Conclusion

This study identified 332 germline variants absent from gnomAD, ExAC, and the 1000 Genomes Project in 55 Malawian patients with DLBCL. These database-absent variants included a higher proportion of predicted loss-of-function consequences and were frequently located in HLA class I loci.

The findings demonstrate limitations in applying global reference databases to underrepresented populations, but they do not establish that the identified variants are Malawian-specific or pathogenic. Larger, well-covered studies with matched Malawian and Southern African controls are required to determine population frequencies and support more reliable germline variant interpretation.

## Data Availability

Drawn from the Kamuzu Central Hospital (KCH) Lymphoma Study, a prospective cohort led by the UNC Project-Malawi Cancer Program since June 2013.

